# Thoracostomy Tube Infections: Prevalence and Associated Clinical Characteristics at a Tertiary Hospital in Northern Tanzania

**DOI:** 10.64898/2026.04.15.26350981

**Authors:** Lucia Damas, Evance S. Rwomurushaka, Moses Sango, Elieshiupendo Niccodem, Theresia E. Mwakyembe, David E. Msuya, Kondo Chilonga

**Affiliations:** Department of General Surgery, KCMC University, Moshi, Tanzania; Department of General Surgery, Kilimanjaro Christian Medical Centre, Moshi, Tanzania; Department of Anatomy and Neuroscience, KCMC University, Moshi, Tanzania; Department of Microbiology and Immunology, KCMC University, Moshi, Tanzania; Department of Microbiology, Kilimanjaro Christian Medical Centre, Moshi, Tanzania

**Keywords:** Thoracostomy tube infection, Chest tube, Antibiotic susceptibility, Northern Tanzania, Surgical site infection

## Abstract

**Background:** Chest tube infection is one of the complications of the tube thoracostomy. Infectious complications may develop in 2% to 25% of patients who undergo thoracotomy tube placement. The use of prophylactic antibiotics to prevent infections associated with thoracostomy tubes remains a subject of debate. Current practices in managing infections related to tube thoracostomy are hindered by the lack of comprehensive and localised data on the microbial profile and their resistance patterns.

**Objective:** To determine the prevalence of thoracostomy tube infections and associated clinical characteristics among patients treated with a thoracostomy tube at KCMC Zonal Referral Hospital.

**Methodology:** Prospective cohort study done at KCMC Zonal Referral Hospital. Include all patients undergoing thoracostomy tube insertion from September 2024 to April 2025.

**Results:** A total of 84 patients underwent tube thoracostomy during the study time. Of these 22 (26.2%) developed SSI. Out of the 22 samples collected, 17 (77.3%) had positive culture results. The most commonly identified pathogens were *Pseudomonas aeruginosa* (41.2%) and *Staphylococcus aureus* (29.4%). The highest overall susceptibility was observed with amikacin, effective against 10 (58.8%) of the tested organisms. The most common resistance was observed against ceftazidime (56.3%) and piperacillin-tazobactam (50.0%). Prolonged chest tube duration (>7 days) was the strongest independent predictor of tube thoracostomy infection.

**Conclusion:** This study revealed a high prevalence of tube thoracostomy infection. Prolonged tube duration and admission to a non-surgical ward care emerge as key risk factors for SSI. These findings underscore the importance of limiting chest tube duration when clinically feasible and ensuring optimal postoperative care environments to minimise the risk of infection.

## INTRODUCTION

A thoracostomy tube is a common procedure in clinical practice that is performed to drain fluid, blood, or air from the pleural cavity (Mullon et al., 2017). It is performed in the setting of life-threatening emergencies or post-operative chest drainage in elective surgery. The common indications include pneumothorax, thoracic empyema, and haemothorax (Pathak et al., 2017). In patients with chest injuries, the majority of patients require thoracostomy tubes. In Tanzania, 38.9% of chest injuries were managed with tube thoracotomy, of which 79.2% were unilateral tube insertions and 20.9% bilateral tube insertions (Mduma et al., 2023).

Thoracostomy tubes are associated with a high complication rate of up to 44.4%, especially when done outside the hospital. These complications are categorised into insertional, positional, and infectious (Jones et al., 2019). Infectious complications account for approximately 1.7% of cases among patients who undergo chest tube insertion for both traumatic and non-traumatic chest conditions (Vilkki and Gunn, 2020). For patients with chest tube insertion due to chest injuries, surgical site infection is the most common complication (Mduma et al., 2023).

Thoracostomy tube infections can manifest as skin infections, empyema, and even necrotising infections. Gram-positive bacteria (GPB) have been reported to be the most frequently isolated bacteria, accounting for 74% of patients with signs of chest tube infections. *Pseudomonas aeruginosa* is a commonly isolated gram-negative bacterium (GNB) (Wright et al., 2021).

Several risk factors have been associated with chest tube insertion infection. These include failure to observe aseptic technique during chest tube insertion, irrational use of antibiotics, comorbidities, smoking, an indication for chest tube insertion, the number of chest tubes inserted, and the duration the patient stays with the chest tube (Hafez et al., 2019).

There is ongoing debate regarding the routine use of antibiotics to reduce infectious complications following tube thoracostomy; however, current evidence is insufficient to support or oppose this practice (Moore et al., 2012). In patients with clinical signs of chest tube-related infections, empirical broad-spectrum antibiotics are initiated after obtaining specimens for culture and sensitivity testing, with subsequent therapy adjusted based on the results.

Antimicrobial resistance (AMR) is currently a global problem. At tertiary hospitals in Northern Tanzania, AMR in bacterial isolates is high (Kumburu et al., 2017). There are currently no local guidelines to guide clinicians in selecting empirical antibiotics for patients with chest tube infections.

Infectious complications may develop in 2% to 25% of patients who undergo thoracostomy tube placement. The infections include pneumonia, empyema, and surgical site infection (cellulitis and necrotising fasciitis) (Kesieme et al., 2011). A recent study at our facility stated that the most frequent complication among patients with chest injuries who had a chest tube inserted was chest tube infection (Mduma et al., 2023).

Old age and diabetes mellitus are associated with increased risk of infectious complications after thoracostomy tube placement (Akiyama et al., 2021; Vilkki and Gunn, 2020). Thoracostomy tube infections are associated with increased antibiotic use, prolonged hospital stays, elevated hospital costs, and mortality. The role of prophylactic antibiotics in preventing infections associated with thoracostomy tubes remains a subject of debate (Platnick et al., 2021).

Current practices in managing infections related to tube thoracostomy are hindered by the lack of comprehensive and localised data on the pathogens involved and their resistance patterns. This knowledge gap can lead to suboptimal treatment regimens, increased morbidity, prolonged hospital stays, and higher health care costs. Moreover, the global rise in antimicrobial resistance highlights the urgent need for reliable data to inform effective treatment protocols.

This study aimed to determine the prevalence, bacterial profile, antimicrobial susceptibility pattern of bacterial isolates, and factors associated with thoracostomy tube infections at a tertiary hospital in Northern Tanzania from September 2024 to April 2025.

The findings of this work are reported in line with the STROBE checklist (Riaz A. et al., 2025).

## METHODOLOGY

### Study design and area

A hospital-based prospective cohort study was conducted at a tertiary hospital in northern Tanzania. This hospital has a 734-bed capacity. It includes departments such as general surgery, medical, oncology, and emergency medicine, among others. The majority of patients with thoracostomy tubes are managed within the named departments. On average, 3 to 6 thoracostomy tube insertions are performed each week, with the majority carried out by the surgical trainees in the general surgery department.

### Study population

This study involved all patients with thoracostomy tubes inserted and admitted at our facility during the study period.

### Sample size and sampling technique

A convenient sampling technique was used. All patients with thoracostomy tubes who met the inclusion criteria and consented to participate in the study were enrolled.

The sample size for this study was 84 respondents, determined using the formula proposed by Fisher. This method incorporated the margin of error (e) and ensured reliable estimation with a conventional confidence level of 95%. A margin of error of 7.8% was selected to achieve a balance between statistical reliability and practical feasibility.

Sample size: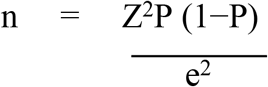

Where by

n = Required sample size

Z = Z-score corresponding to the desired confidence level (e.g., 1.96 for 95% confidence)

P = estimated proportion of patients with surgical site infection following thoracostomy tube insertion

e = margin of error (usually set at 7.8% or 0.078)

According to Naing et al. (2022), while well-funded or large-scale studies often target a precision of 2–3%, small-scale or student-led studies can reasonably adopt a wider margin of 5–10%, particularly when estimating prevalence within a 10–90% range (Naing et al., 2022). A margin of error of 7.8% is well within the commonly accepted range for small-scale or resource-limited studies, allowing for reliable interpretation of prevalence estimates while maintaining feasibility given available resources. Similarly, Das et al. (2020) emphasize that smaller margins of error require significantly larger sample sizes, which may be impractical in resource-constrained settings (Das et al., 2020). Therefore, the chosen precision level is not only statistically valid but also contextually appropriate for a baseline epidemiological investigation at our facility.

Estimated Proportion from prior studies

- P-Proportion of patients with TTI predicted in a study done at our facility (14%) (Mduma et al., 2023).
- Therefore, P=0.14 (14%).

Sample size:

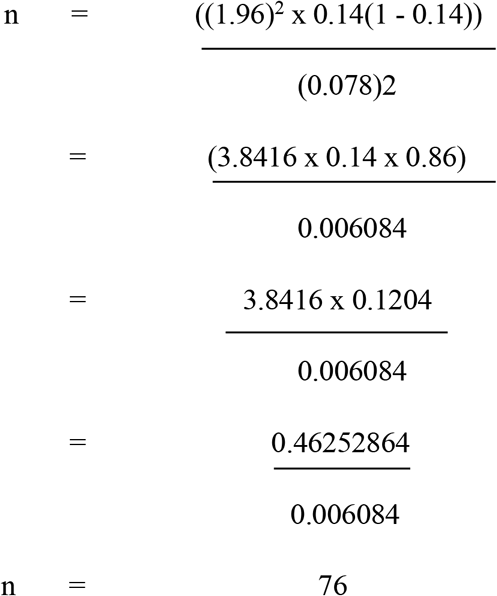

Adjust for Attrition: Addition of 10% allowance for potential non-responses or exclusion.

n =76 + (76 × 10% or 0.1).

n = 76 + 7.6

n = 83.6 or 84

A total of 84 patients were recruited to ensure adequate precision in estimating the prevalence of surgical site infections and to account for potential non-response or loss to follow-up.

### Dependent variables

Thoracostomy tube infection, bacterial isolates, and antimicrobial susceptibility.

### Independent variables

Age, sex, indication for chest tube insertion, place where the chest tube was inserted (EMD, MOT, WARD), ward of admission, history of antibiotic use, number of chest tubes inserted, duration of the chest tube, and comorbidities.

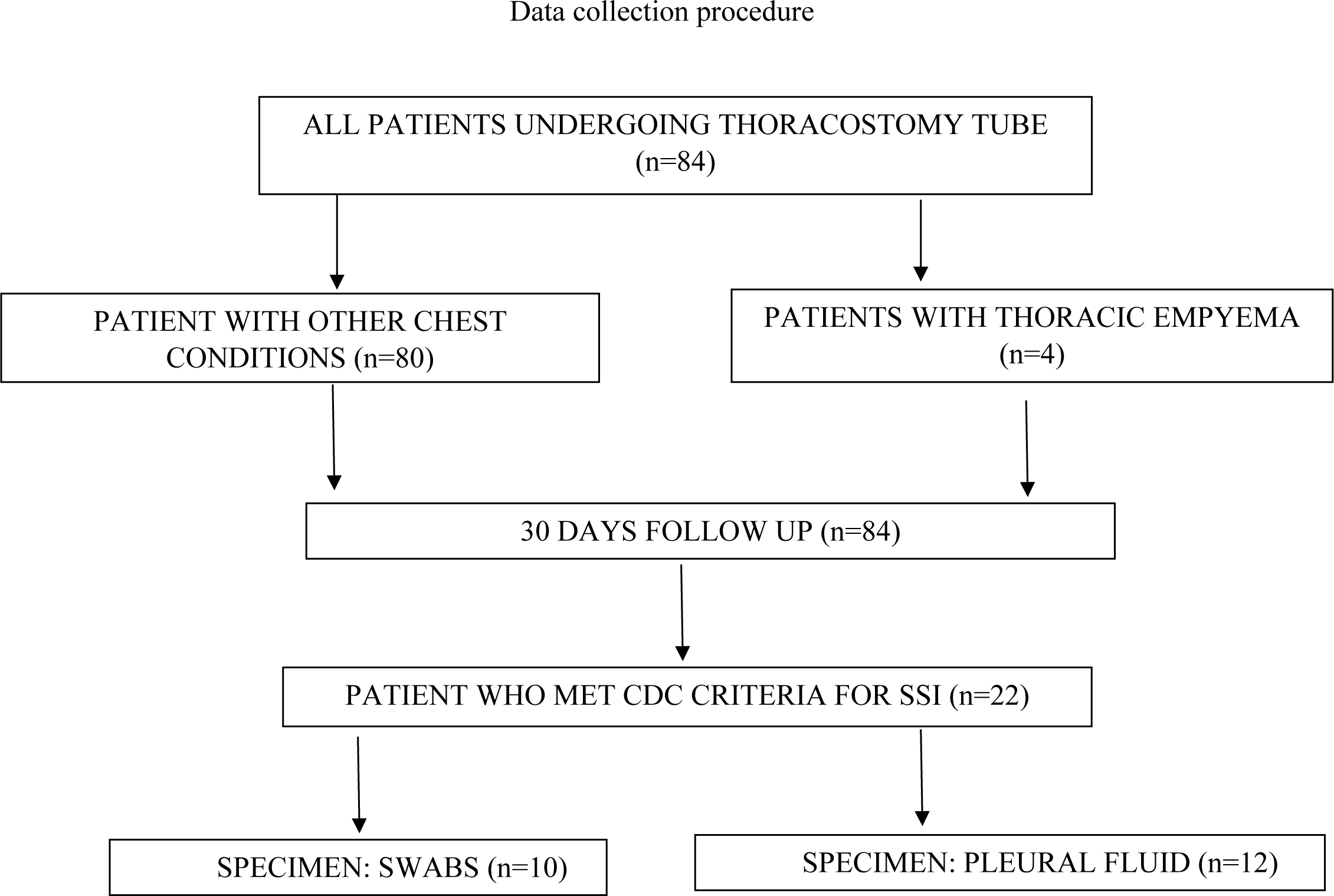

### Sample collection procedure

Patients who developed signs of infection at the thoracostomy site were identified. A pus swab was taken from the deepest part of the wound in patients with discharge, and pleural fluid was aspirated from the pleural cavity using a syringe in patients draining purulent fluid through the chest tube. The swabs were collected in Amies agar media and immediately transported to the laboratory for processing. For patients with fluid, 2mL of the fluid was collected in a vacuum container and immediately transported to the laboratory for processing.

### Data collection tool

All collected data were filled into an Excel data sheet.

### Data management and analysis plan

All data were analysed by the Statistical Package for Social Sciences (SPSS) version 23. Descriptive statistics were summarised using frequency and percentage for categorical variables, and continuous variables were summarised using measures of central tendency and their respective measures of dispersion. Logistic regression model test was used to measure the associations, and a p-value of < 0.05 was considered statistically significant.

### Ethical considerations

Ethical clearance was granted from the institutional review board. Permission to conduct research was obtained from the Executive Director of the hospital from which data was collected. Patient written consent or assent to take part in the study was sought. Confidentiality was maintained in all stages of handling data. Patients were given special identification. Patients diagnosed with tube thoracostomy infection with positive culture and sensitivity results were treated with the antibiotics according to susceptibility results.

## RESULTS

A total of 84 patients underwent tube thoracostomy during the study time. All patients consented to the study, all were followed up within 30 days post-tube thoracostomy, and all were available for the final analysis.

### Clinical characteristics of patients treated with tube thoracostomy from September 2024 to April 2025

The median age of the study population was 41.5 years (IQR: 26.25–59.25), with the age group ≥46 years 40 (47.6%). Males were predominant (56.0%). The majority of chest tubes were inserted in emergency situations (77.4%). Half of the study population was admitted to wards outside general surgery wards (Table 1). Over half of the patients (54.8%) had thoracostomy tubes in place for less than 7 days. Right-sided chest tubes were inserted in 47.6% of patients, while left-sided tubes accounted for 34.9 %, and the remainder underwent bilateral thoracostomy tube insertion.

**Table 1:** Clinical characteristics of patients treated with tube thoracostomy from September 2024 to April 2025 (N=84)

| Characteristics | Frequency (%) |
| --- | --- |
| Age |  |
| $\leq 25$ | 20(23.8) |
| 26-45 | 24(28.6) |
| $\geq 46$ | 40(47.6) |
| Median (IQR) | 41.5(26.25-59.25) |
| Sex |  |
| Female | 37(44.0) |
| Male | 47(56.0) |
| Comorbidity |  |
| Yes | 55(65.5) |
| No | 29(34.5) |
| Urgency |  |
| Emergency | 65(77.4) |
| Elective | 19(22.6) |
| Number of days chest tube stay |  |
| $\leq 7$ | 46(54.8) |
| $> 7$ | 38(45.2) |
| Median (IQR) | 7(4-11) |
| Number of chest tubes inserted |  |
| One | 69(82.1) |
| Two | 15(17.9) |
| Side chest tube inserted |  |
| Right | 40(47.6) |
| Left | 29(34.5) |
| Both | 15(17.9) |
| Admission ward |  |
| Surgical Ward | 42(50.0) |
| Non-Surgical Ward | 42(50.0) |

### The Prevalence of surgical site infection among patients with a tube thoracostomy from September 2024 to April 2025

Among 84 patients who underwent chest tube insertion, 22 (26.2%) developed SSI. Of these, 17 (77.3%) had positive culture results, and 5 (22.7%) had no identifiable microbial isolates. The SSI cases occurred in patients with chest tube stay >7 days (77.3%). Inpatient procedures were reported in 14 (63.6%) of infections (Table 2).

**Table 2:** Characteristics of patients who developed SSI among patients with tube thoracostomy from September 2024 to April 2025 (N=22).

| Characteristics | Frequency (%) |
| --- | --- |
| Age |  |
| ≤25 | 6(27.3) |
| 26-45 | 7(31.8) |
| ≥46 | 9(40.9) |
| Sex |  |
| Female | 11(50.0) |
| Male | 11(50.0) |
| Comorbidities |  |
| Yes | 21(95.5) |
| No | 1(4.5) |
| Urgency |  |
| Emergency | 15(68.2) |
| Elective | 7(31.8) |
| Number of days chest tube stay |  |
| ≤7 | 5(22.7) |
| >7 | 17(77.3) |
| Number of chest tubes inserted |  |
| One | 20(90.9) |
| Two | 2(9.1) |
| Admission ward |  |
| General surgery ward | 5(22.7) |
| Non-general surgery ward | 17(77.3) |
| Culture results |  |
| Positive | 17(77.3) |
| Negative | 5(22.7) |
| Indication based on causes |  |
| Chest trauma | 4(18.2) |
| Non chest trauma | 18(81.8) |
| Patient on antibiotics |  |
| No | 2(9.1) |
| Yes | 20(90.9) |

### The bacterial profile among patients with tube thoracostomy infection from September 2024 to April 2025

A total of 22 patients who developed SSI had specimens submitted for culture. Of the submitted specimens, 12 (54.5%) were fluid samples, while 10 (45.5%) were wound swabs. Culture results were positive in 17 (77.3%) of the cases. Among the isolates, the most commonly identified pathogens were *Pseudomonas aeruginosa* (41.2%) and *Staphylococcus aureus* (29.4%) (Table 3). Most isolates were recovered from patients managed in non-surgical wards, including 6 (85.7%) of *Pseudomonas aeruginosa* and 4 (80.0%) of *Staphylococcus aureus*. Infections among patients with non-traumatic causes contributed the majority of isolates, including 100.0% of *Citrobacter freundii* and *Klebsiella pneumoniae*, 85.7% of *Pseudomonas aeruginosa*, and 80.0% of *Staphylococcus aureus* (Table 4).

**Figure 1.**
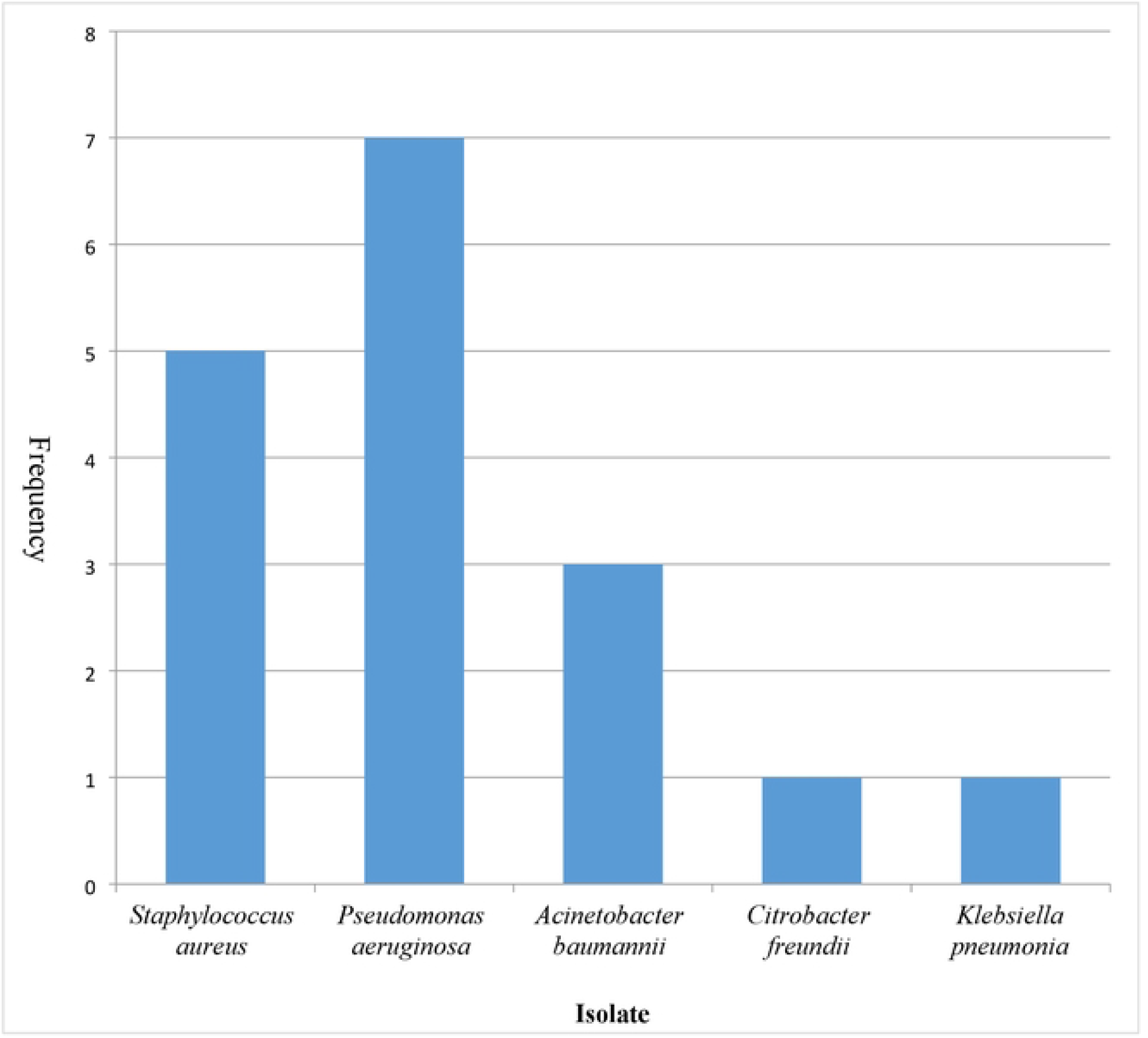
Bacterial profile among patients with tube thoracostomy infection (N=17)

**Table 3:** Distribution of isolates among patients with tube thoracostomy infection.

|  | *A.baumannii*<br>n=3 | *C. freundii*<br>n=1 | *K.pneumoniae*<br>n=1 | *P. aeruginosa*<br>n=7 | *S.aureus*<br>n=5 |
| --- | --- | --- | --- | --- | --- |
| Characteristics | Frequency (%) | Frequency (%) | Frequency (%) | Frequency (%) | Frequency (%) |
| ward patient cared |  |  |  |  |  |
| Surgical Ward | 1(33.3) | 1(100) | 0(0) | 1(14.3) | 1(20.0) |
| Non-Surgical Ward | 2(66.7) | 0(0) | 1(100) | 6(85.7) | 4(80.0) |
| Indication of TT |  |  |  |  |  |
| Chest trauma | 1(33.3) | 0(0) | 0(0) | 1(14.3) | 1(20.0) |
| Non-chest trauma | 2(66.7) | 1(100) | 1(100) | 6(85.7) | 4(80.0) |

**Table 4:** Antimicrobial resistance to isolated organisms.

| % Resistance |  |  |  |  |  |  |  |  |  |  |  |  |  |  |  |  |
| --- | --- | --- | --- | --- | --- | --- | --- | --- | --- | --- | --- | --- | --- | --- | --- | --- |
| Bacterial isolate | n | CRO | CIP | FE | CN | ME | CTX | SXT | TZP | CZ | AK | ER | CHL | TC | AMC | AMP |
| *A. baumannii* | 3 | 13 | 9 | 9 | 9 | 13 | 4 | 13 | 13 | 13 | 4 | - | - | - | - | - |
| *C. freundii* | 1 | 20 | - | - | - | - | - | - | 20 | 20 | - | - | - | - | 20 | 20 |
| *K. pneumonia* | 1 | 20 | 20 | - | 20 | - | - | 20 | - | 20 | - | - | - | - | - | - |
| *P. aeruginosa* | 7 | - | 20 | 7 | - | 13 | - | - | 27 | 27 | - | - | - | - | - | - |
| *S. aureus* | 5 | - | 10 | - | - | - | - | 20 | - | - | 10 | 30 | 10 | 10 | - | - |

KEY: CRO: Ceftriaxone, TZP: Piperacillin/Tazobactam, AMC: Amoxicillin/Clavulanic acid, CN: Gentamicin, CIP: Ciprofloxacin, ME: Meropenem, SXT: Trimethoprim Sulfamethoxazole, CZ: Ceftazidime, ER: Erythromycin, AMP: Ampicillin, CHL: Chloramphenicol, FE: Cefepime, CTX: Cefotaxime, AK: Amikacin, TC: Tetracycline

### Antimicrobial susceptibility pattern among patients with tube thoracostomy infection from September 2024 to April 2025

The antimicrobial resistance reveals a concerning pattern of multidrug resistance among the 17 bacterial isolates, especially among Gram-negative organisms. *Acinetobacter baumannii* demonstrated the highest level of resistance, showing 100% resistance to several antibiotics, including ceftriaxone, meropenem, trimethoprim-sulfamethoxazole, piperacillin-tazobactam, and cefazolin. It demonstrated moderate resistance to ciprofloxacin, cefepime, gentamicin, and amikacin. *Citrobacter freundii* showed complete resistance to ceftriaxone, piperacillin-tazobactam, cefazolin, amoxicillin-clavulanic acid, and ampicillin. *Klebsiella pneumoniae* exhibited moderate resistance to several antibiotics, including ceftriaxone, ciprofloxacin, Gentamicin, and trimethoprim-sulfamethoxazole. *Pseudomonas aeruginosa* showed notable resistance to piperacillin-tazobactam and cefazolin, with lower resistance to ciprofloxacin, cefepime, and meropenem. In contrast, *Staphylococcus aureus* displayed lower overall resistance, with the highest resistance observed to erythromycin (30%) and moderate resistance to trimethoprim-sulfamethoxazole, tetracycline, and chloramphenicol (Table 4)

### Factors associated with tube thoracostomy infection among patients with tube thoracostomy from September 2024 to April 2025

The chi-square analysis revealed that several variables had a statistically significant association with the development of SSI. Prolonged chest tube duration (>7 days) was significantly associated with SSI (χ^2^ = 12.347, p < 0.001), with 77.3% of infections occurring in this group. Similarly, the type of ward where patients were managed was significant (χ^2^ = 8.868, p = 0.003), with most infections (77.3%) occurring in non-surgical wards. The underlying cause for tube insertion also showed a strong association (χ^2^ = 7.441, p = 0.006), where 81.8% of SSI cases were related to non-traumatic conditions. (Table 5) Multivariable logistic regression identified prolonged chest tube duration (>7 days) as the strongest independent predictor of SSI, having 1.39 times higher odds of developing infection compared to those with ≤7 days of tube stay (aOR = 1.39, 95% CI: 1.18–1.64, p < 0.001) (table 6).

**Table 5:** Predictors’ association with SSI.

| Characteristics | Developed SSI |  |  | X<sup>2</sup>(p-value) |
| --- | --- | --- | --- | --- |
|  | n (%) | No n (%) | Yes n (%) |  |
| Age |  |  |  | .542(.763) |
| $\leq 25$ | 20(23.8) | 14(22.6) | 6(27.3) | |
| 26-45 | 24(28.6) | 17(27.4) | 7(31.8) |  |
| $\geq 46$ | 40(47.6) | 31(50.0) | 9(40.9) | |
| Sex |  |  |  | .428(.513) |
| Female | 37(44.0) | 26(41.9) | 11(50.0) |  |
| Male | 47(56.0) | 36(58.1) | 11(50.0) |  |
| Urgency | | | | 1.441(.230) \$ |
| Elective | 19(22.6) | 12(19.4) | 7(31.8) |  |
| Emergency | 65(77.4) | 50(80.6) | 15(68.2) |  |
| Duration of Tube Stay |  |  |  | 12.347(<.001) * |
| $\leq 7$ | 46(54.8) | 41(66.1) | 5(22.7) | |
| $> 7$ | 38(45.2) | 21(33.9) | 17(77.3) | |
| Number of chest tubes inserted | | | | 1.562(.211) \$ |
| One | 69(82.1) | 49(79.0) | 20(90.9) |  |
| Two | 15(17.9) | 13(21.0) | 2(9.1) |  |
| Side chest tube inserted | | | | 1.612(.447) \$ |
| Right | 40(47.6) | 28(45.2) | 12(54.5) |  |
| Left | 29(34.5) | 21(33.9) | 8(36.4) |  |
| Both | 15(17.9) | 13(21.0) | 2(9.1) |  |
| The ward patient has been cared |  |  |  | 8.868(.003) * |
| Surgical Ward | 42(50.0) | 37(59.7) | 5(22.7) |  |
| Non-Surgical Ward | 42(50.0) | 25(40.3) | 17(77.3) |  |
| Indication based on causes |  |  |  | 7.441(.006) * |
| Chest trauma | 36(42.9) | 32(51.6) | 4(18.2) |  |
| Non-chest trauma | 48(57.1) | 30(48.4) | 18(81.8) |  |
| Comorbidities |  |  |  | 11.850(.001) * |
| No | 29(34.5) | 28(96.6) | 1(3.4) |  |
| Yes | 55(65.5) | 34(61.8) | 21(38.2) |  |
| Patient on antibiotics |  |  |  | .171(.680) |
| No | 6(7.1) | 4(6.5) | 2(9.1) |  |
| Yes | 78(92.9) | 58(93.5) | 20(90.9) |  |

\$=to be included in univariate analysis; Chi-Square; \* Significance

**Table 6:** Multivariable Logistic Regression for Factors Associated with SSI.

| Characteristics | Developed SSI |  |  |  |
| --- | --- | --- | --- | --- |
|  | cOR(95%CI) | p-value | aOR(95%CI) | p-value |
| Urgency |  |  |  |  |
| Elective | ref |  | ref |  |
| Emergency | 1.23(1.18-1.43) | <.001** | .96(.74-1.23) | .959 |
| Duration of Tube Stay |  |  |  |  |
| ≤7 | ref |  | ref |  |
| >7 | 1.40(1.17-1.68) | <.001** | 1.39(1.18-1.64) | <.001* |
| The ward patient has been cared |  |  |  |  |
| Surgical Ward | ref |  | ref |  |
| Non-Surgical Ward | 1.33(1.11-1.59) | .002* | 1.05(.84-1.31) | .685 |
| Indication based on causes |  |  |  |  |
| Chest trauma | ref |  | ref |  |
| Non-chest trauma | 1.30(1.09-1.5) | .003* | .98(.75-1.29) | .887 |
| History of cigarette smoking |  |  |  |  |
| No | ref |  | ref |  |
| Yes | .89(.72-1.09) | .224 | .94(.78-1.13) | .489 |

cOR=Crude Odds Ratio; aOR= Adjusted Odds Ratio; \*Statistically Significant

## DISCUSSION

A thoracostomy tube is a common procedure in clinical practice, performed to drain fluid, blood, or air from the pleural cavity. Chest tube infection is a common complication associated with this procedure. Infectious complications may develop in 2% to 25% of patients who undergo thoracostomy tube placement.

The main objective of this study was to determine the prevalence of tube thoracostomy infections and associated clinical characteristics among patients treated with a thoracostomy tube at a tertiary hospital in northern Tanzania from September 2024 to April 2025. A total of 84 patients had chest tubes inserted during the study period; all consented to participate in the study and were available for the final analysis.

The prevalence of thoracostomy tube infection was 26.2%. These infections were highest in patients admitted to outside general surgery wards, patients who stayed with a chest tube for more than 7 days, and those who had tubes due to other chest diseases apart from chest trauma. The prevalence in this study was higher compared to studies in Finland, the USA, and Bangladesh (Akiyama et al., 2021; Vilkki and Gunn, 2020). It was also higher than the rate reported at the same facility two years earlier (Mduma et al., 2023). The difference in study population explains the difference in prevalence.

The bacterial spectrum observed in this study was predominantly *Pseudomonas aeruginosa* (41.2%) and *Staphylococcus aureus* (29.4%). These findings are consistent with a previous study at our facility, which investigated surgical site infections, in which *Pseudomonas aeuruginosa* and *Staphylococcus aureus* were the predominant isolates. This highlights the predominance of these organisms in post-procedural infections (Kumburu et al., 2017). This prevalence is comparable to that found at Muhimbili National Hospital (Massaga and Mchembe, 2010).

These results differ from a study conducted in Japan, where *Cutibacterium acnes* was the most commonly isolated organism, while *Staphylococcus epidermidis* and *Staphylococcus capitis* were identified less frequently (Akiyama et al., 2021). *Staphylococcus aureus*, being a part of normal flora, is more likely to invade the surgical site or contaminate the specimen during collection. Although the sample size was small in the current study, the geographical area and underlying pathology may have contributed to the similarities and variations observed across studies.

The world is currently advocating for the use of antibiotics based on culture and sensitivity results, as antimicrobial resistance is rising despite advancements in the pharmaceutical industry. In the current study, amikacin demonstrated the highest effectiveness, with 10 (58.8%) of the tested organisms showing susceptibility, followed by ciprofloxacin, which was effective in 9 (52.9%) organisms. Resistance testing revealed high levels of antimicrobial resistance among isolated pathogens.

The most common resistance was observed against ceftazidime (56.3%), followed by piperacillin-tazobactam (50.0%), and ciprofloxacin (43.8%). The resistance to ciprofloxacin was also observed in the previous study at our facility and in Ethiopia (Alelign et al., 2022; Kumburu et al., 2017). Notable ciprofloxacin resistance could be attributed to its pattern of use, similar to what has been reported with ceftriaxone in previous studies (Kumburu et al., 2017; Yamauchi et al., 2013). Ceftriaxone, the first-line therapy in surgical patients at our facility, was noted to have the lowest susceptibility and moderate resistance. This is alarming, given the fact that ceftriaxone is affordable to the majority of patients.

The number of days the patient stayed with a chest tube has been noted to be associated with the tube thoracostomy infection. A longer duration of tube placement was associated with an increased risk of tube thoracostomy infection. In the current study, a statistically significant relationship was observed between the duration of tube thoracostomy and the occurrence of infection. Patients with chest tubes for more than 7 days have higher odds of tube thoracostomy infection compared to those who had a chest tube for less than 7 days. This was similar to a study in Canada (Oldfield et al., 2009) and another study in Egypt (Hafez et al., 2019). Though a tube thoracostomy is therapeutically beneficial, prolonged placement can increase the risk of infection.

Emergency chest tube insertions are often performed under suboptimal conditions, increasing the risk of contamination. Inadequate dressing change, poor hygiene practices, and delayed recognition of site redness or discharge are associated with higher infection rates (Freeman et al., 2022). Biofilms on the surface of a chest tube can shield bacteria from antibiotics and the host immune system, facilitating chronic infections (Deshpande et al., 2024). Similar to our study, patients who underwent chest tube insertion under emergency conditions had a higher frequency of chest tube infection. In our study, patients with a thoracostomy tube admitted to the surgical ward had a lower infection rate compared to those admitted to non-surgical wards. This may indicate better thoracostomy tube care in the surgical ward, with improved hygiene and frequent dressing changes, which disrupt the biofilm on the thoracostomy tube, hence decreasing infection rates.

## CONCLUSION

This study revealed that the prevalence of tube thoracostomy infection is high. Prolonged tube duration and non-surgical ward care emerge as key risk factors for SSI. Timely removal of chest tubes is vital in minimizing the risk of infection.

## Data Availability

Data collected for this study is present in an excel sheet held by the corresponding author, and will be presented upon request by the editor of this journal.

## Notes

### Competing Interest Statement

The authors have declared no competing interest.

### Funding Statement

The author(s) received no specific funding for this work.

### Author Declarations

Kilimanjaro Christian Medical University College (KCMUCo) Clinical Research and Ethics Committee

